# Accuracy and performance of the WHO 2015 and WHO 2019 HIV testing strategies across epidemic settings

**DOI:** 10.1101/2021.03.31.21254700

**Authors:** Jeffrey W Eaton, Anita Sands, Magdalena Barr-DiChiara, Muhammad S Jamil, Rachel Baggaley, Beth A Tippett Barr, Thokozani Kalua, Andreas Jahn, Mathieu Maheu-Giroux, Cheryl C Johnson

## Abstract

**Background:** WHO 2019 HIV testing guidelines recommended a standard HIV testing strategy consisting of three consecutively HIV-reactive test results on serology assays to diagnose HIV infection. National HIV programmes in high prevalence settings currently using the strategy consisting of only two consecutive HIV-reactive tests should consider when to implement the new guideline recommendations.

**Methods and Findings:** We implemented a probability model to simulate outcomes of WHO 2019 and the two strategies recommended by WHO 2015 guidelines on HIV testing services. Each assay in the strategy was assumed independently 99% sensitivity and 98% specificity, the minimal thresholds required for WHO prequalification. For each strategy and positivity ranging 20% to 0.2%, we calculated the number of false-negative, false-positive, and inconclusive results; positive and negative predictive value (PPV, NPV); number of each assay used, and testing programme costs. We found that the NPV was above 99.9% for all scenarios modelled. Under the WHO 2015 two-test strategy, the PPV was below the 99% target threshold when positivity fell below 5%. For the WHO 2019 strategy, the PPV was above 99% for all positivity levels. The number reported ‘inconclusive’ was higher under the WHO 2019 strategy. Implementing the WHO 2019 testing strategy in Malawi, would require around 89,000 A3 tests in 2021, compared to 175,000 A2 tests and over 4.5 million A1 tests per year. The incremental cost of the WHO 2019 strategy was less than 2% in 2021 and declined to 0.9% in 2025.

**Conclusions:** As positivity among persons testing for HIV reduces below 5% in nearly all settings, implementation of the WHO 2019 testing strategy will ensure that positive predictive value remains above the 99% target threshold, averting misdiagnoses and ART initiations among HIV uninfected people. The incremental cost of implementing the WHO 2019 HIV testing strategy compared to the two-test strategy is negligible because the third assay accounts for a small and diminishing share of total HIV tests.

## Introduction

Providing accurate, timely, and affordable HIV diagnosis at the point of care is a critical first step towards delivering HIV treatment and prevention services. To establish HIV diagnosis, the World Health Organization (WHO) recommends HIV testing strategies using multiple HIV serology assays including rapid diagnostic tests (RDTs) and enzyme-immunoassays (EIAs) [1,2]. Each assay should have a sensitivity of greater than or equal to 99% (the assay is reactive for at least 99 out of 100 truly HIV positive specimens) and specificity of greater than or equal to 98% (the assay is non-reactive for at least 98 out of 100 truly HIV negative specimens), the minimum thresholds for approval through the WHO prequalification procedure [3,4].

To ensure accurate diagnosis in all settings, WHO has previously recommended differentiated HIV testing strategies according to the prevalence of HIV in the population being tested [2,4]. In ‘high’ HIV prevalence populations with prevalence above 5%, two consecutively HIV-reactive test results were recommended to diagnose HIV; when prevalence was below 5%, three consecutively HIV-reactive test results were recommended. This threshold at 5% was established to ensure that the combination of assays utilized in a testing strategy have at least 99% positive predictive value (PPV)—that is at least 99 out of every 100 individuals classified as HIV positive are truly HIV positive, or fewer than 1 false-positive per 100 HIV positive individuals tested— when assays attain the minimum 98% specificity requirement [4].

When choosing a testing strategy, HIV programmes have typically used national HIV population prevalence (greater or less than 5%) as a proxy to guide whether to implement a ‘two-test’ or ‘three-test’ strategy. However, as awareness of HIV status has reached high levels, positivity amongst persons presenting for HIV testing services (HTS) has declined steeply in recent years. Even in the highest HIV prevalence settings in southern and eastern Africa, the positivity is below the 5% threshold and expected to further decline in future given high ART coverage and estimated low HIV incidence [5]. A 2018 policy review found that only 25% of national HIV testing guidelines were fully adherent with WHO recommendations on HIV testing strategies [6]. Despite declining HIV positivity, few programmes have transitioned from using two HIV-reactive test results to three HIV-reactive test results to maintain high PPV.

Responsive to declining positivity and the challenges implementing differentiated guideline recommendations, the 2019 revision of the *WHO Consolidated Guidelines on HIV Testing Services* recommended a single ‘standard’ HIV testing strategy for all settings in which HIV testing is conducted. The WHO 2019 testing strategy recommended HIV-reactive results from three consecutive serology assays and simplified the steps for adjudicating cases with discrepant results on serially conducted tests (Figure 1). National HIV programmes should now consider when and how to implement new recommendations to transition from a ‘two-test’ to the ‘three-test’ testing strategy, including the potential implications for accuracy and misdiagnosis, and programme and commodities costs.

**Figure 1.**
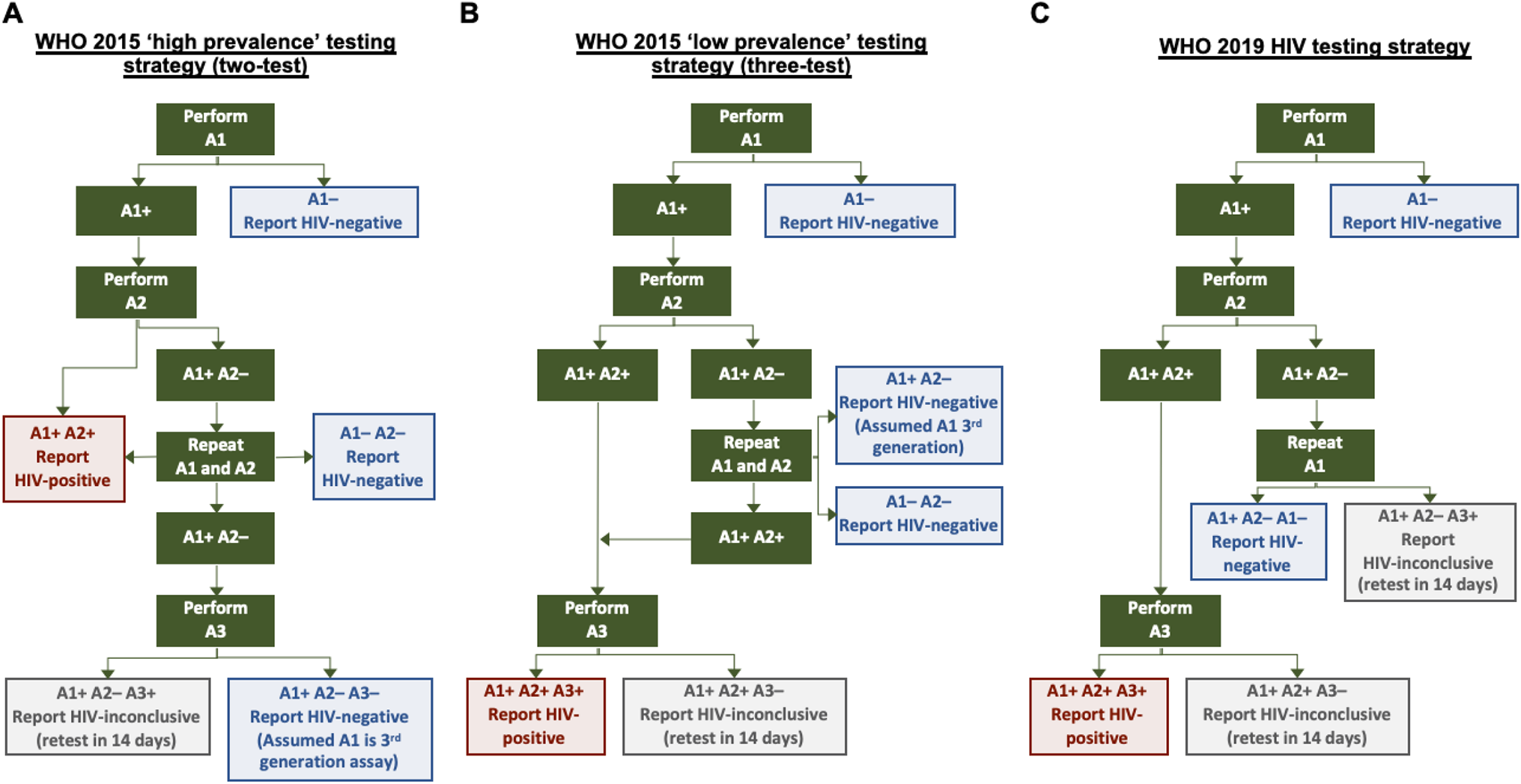
HIV testing strategies modelled. (A) and (B) are redrawn following *WHO Consolidated Guidelines on HIV Testing Services 2015* Figures 7.2 and 7.3 [4]. (A) and (B) assume a 3^rd^ generation A1 assay is used. (C) is redrawn following *WHO Consolidated Guidelines on HIV Testing Services 2019* Figure 8.3 [7].

To support decision making, we analysed the performance of the WHO 2019 standard HIV testing strategy compared to the previous two-test and three-test testing strategies with respect to PPV, negative predictive value (NPV), and the number of HIV-inconclusive results returned at a range of HIV positivity values. Second, we calculated cost and commodity considerations of the respective testing strategies using epidemiologic and HIV testing programme data from Malawi as an example case study.

## Methods

### Model

We implemented a probability model to deterministically simulate the expected outcomes of the WHO 2019 HTS Guidelines and the two testing strategies recommended by WHO 2015 HTS Guidelines, including stipulated repetition of one or more assays in the case of discrepant test results (Figure 1) [4,7]. We refer to the ‘high’ and ‘low’ prevalence testing strategies as the ‘two-test’ and ‘three-test’ strategies, respectively. The numbers refer to the sequential assays required for an HIV-positive diagnosis, and not the overall number of assays needed in the respective strategy, as the two-test testing strategy also requires a third assay to be available in the event of discrepant results (Figure 1A) [4].

Each assay in the testing strategy was assumed to be independent and perform with 99% sensitivity and 98% specificity, the minimum performance requirements for WHO prequalification [7,8]. WHO recommends that any initially discrepant A1+/A2– cases be repeated immediately using assay 1 (A1). We assumed that repetition of A1 will resolve the discrepancy in 80% of cases (e.g. with presumed ‘use error’ or other random error resulting in initially discrepant results), while the A1+/A2–/A1+ discrepancy persisted 20% of the time (due to false reactivity for A1+ for truly HIV negative persons or low antibody levels not detected with A2 for truly HIV positive persons). Case that were twice discrepant (initial: A1+ / A2–; repeat: A1+ / A2–) under the WHO 2015 three-test testing strategy were classified as ‘HIV negative’. (This outcome does not occur under the WHO 2019 strategy because A2 is not repeated. Such cases A1+ / A2– / A1+ are reported HIV-inconclusive after repeat A1+ and recommended to retest after 14 days).

Input parameters to the model were: (1) Number individuals tested for HIV, (2) proportion truly HIV positive among those tested, (3) WHO HIV testing strategy (two-test or three-test), (4) sensitivity of each assay (assumed 99%), (5) specificity of each assay (assumed 98%), (6) probability that repetition of false-reactive A1 (A1+ for a truly HIV-negative person) is non-reactive upon repetition of A1 (assumed 80%), and (7) HIV testing costs (including commodities, site overheads, staff salaries, and other).

### Outputs and cost analysis

The model produced the following outcomes: the absolute number of individuals ruled HIV-positive, HIV-negative, and HIV-inconclusive (who are asked to return after 14 days to undergo testing again), observed HIV-positivity amongst individuals presenting for HTS, number false-positive and number false-negative diagnoses, positive and negative predictive values, number of A1, A2, and A3 products utilized, and estimated total HIV testing costs.

Costs were indicative of current HTS programme costs in many low- and middle-income (LMIC) settings and were ‘fully loaded’ costs including consumables, staff, facilities, and programme management. We assumed an average service delivery cost of US$2.00 per client tested plus the unit cost of each assay used in the algorithm. The unit cost reported in the WHO Global Price Reporting for a typical product used as A1 was around $0.80 per test compared to around $1.60 for a typical product used as A2 [9]. Supply chain monitoring data from Malawi indicate that around 95% of A1 consumables are used for client testing with the remaining 5% for other purposes such as quality assurance, proficiency testing, and loss. For A2 products, used only after an HIV-reactive A1 result, around 70% are used for client testing and 30% for other purposes [10]. Accounting for these factors and other per-test consumables and staff expenditures, we assumed US$1.30 for each A1 utilized, $2.30 for each A2 utilized, and US$2.50 per A3 utilized. Taken together, the cost was around $3.50 per HIV-negative client tested and $5.60 to $8.10 per HIV-positive person tested. For cases initially reported as ‘HIV-inconclusive’, we accounted for testing commodities and cost for retesting after 14 days, but did not count the HIV status reported at repeat testing in calculations for number classified positive, negative, PPV, or NPV because the primary objective of the HIV testing strategy is to return the accurate HIV testing result at the clinical encounter.

### Analyses

We conducted two analyses. First, we simulated the expected HIV testing outcomes per 100,000 individuals tested using either the WHO 2019, WHO 2015 two-test or WHO 2015 three-test strategies for true positivity ranging from 20% to 0.2%. We assessed (1) whether each test strategy achieved above the 99% threshold for PPV and NPV when assuming assays performed at the minimum level required for WHO prequalification, (2) the number of HIV-inconclusive results returned, and (3) the number of tests and total HTS cost.

Secondly, we used HIV epidemic estimates and projections and HIV testing programme data from Malawi to understand the consequences of changes in HIV testing scale and positivity over time on expected HIV testing programme outcomes.

We used 2020 national HIV estimates for Malawi submitted to UNAIDS to extract estimates for the adult (15+) population over the period 2000 to 2025 for the indicators: number of people living with HIV (PLHIV), number aware of HIV status and number on ART, and number of HIV tests conducted, number of persons testing positive for HIV, and number persons diagnosed HIV positive for the first time. Estimates and projections for the number of PLHIV and number on ART were estimated by the Malawi national HIV estimates working group using the Spectrum and EPP models [11,12]. HIV testing programme outcomes over the period 2000 to 2019 were estimated from household survey and HIV testing programme data input to the Shiny90 model [13]. Projections for the period 2020 through 2025 assumed that the 2019 rates of HIV testing by age, sex, HIV status, and ART status would continue into the future. We input the estimates for the annual number of tests conducted by HIV status into the HIV testing strategy model to estimate the expected HIV testing programme outcomes had the two-test or WHO 2019 testing strategy been used, and assuming assay performance at the minimum level to attain WHO prequalification.

The Malawi HIV testing programme has implemented the high prevalence (two-test) testing strategy for HIV diagnosis, but the simulated results do not represent actual HIV testing programme outcomes for Malawi in at least two ways. Firstly, HIV testing programme data from Malawi suggest improving accuracy over time following a programme of training and quality assurance. Secondly, since 2011 Malawi has implemented retesting for verification of HIV status of all HIV-positive persons before ART initiation [14], which is not included in our simulation.

## Results

### Outcomes per 100,000 tested for range of HIV testing positivity

Table 1 summarizes model outcomes per 100,000 persons tested at 10%, 5%, 1%, and 0.5% true positivity amongst persons tested for HIV. The expected number of false-positive classifications is substantially lower with the three-test and WHO 2019 testing strategies at fewer than 1 false HIV-positive per 100,000 individuals tested compared to around 45 false-positives per 100,000 individuals tested with the two-test strategy and does not vary substantially with positivity (Table 1, Figure 2A). In contrast, the expected number of false-negative classifications was proportional to the positivity from 100 false-negatives per 100,000 individuals tested at 10% positivity to 10 false-negative per 100,000 individuals tested at 1% positivity with the two-test strategy. The three-test or WHO 2019 testing strategy does not affect the expected number of false-negative classifications.

**Table 1.** HIV testing strategy outcomes per 100,000 tested for 10%, 5%, 1%, and 0.5% true positivity among clients presenting for HIV testing

|  | 10% positivity |  |  | 5% positivity |  |  |
| --- | --- | --- | --- | --- | --- | --- |
|  | 2-test | 3-test | WHO 2019 | 2-test | 3-test | WHO 2019 |
| HIV-negative classifications | 90,022 | 90,049 | 89,712 | 94,968 | 94,985 | 94,640 |
| HIV-positive classifications | 9922 | 9781 | 9704 | 4985 | 4891 | 4852 |
| HIV-inconclusive | 55 | 170 | 584 | 47 | 124 | 508 |
| Observed positivity | 9.93% | 9.80% | 9.76% | 4.99% | 4.90% | 4.88% |
| False HIV-positive | 43.1 | 0.86 | 0.72 | 45.4 | 0.91 | 0.76 |
| False HIV-negative | 100.4 | 119.8 | 101.0 | 50.2 | 59.9 | 50.5 |
| PPV of entire testing strategy | 99.6% | >99.9% | >99.9% | 99.1% | >99.9% | >99.9% |
| NPV of entire testing strategy | 99.9% | 99.9% | 99.9% | 99.9% | 99.9% | 99.9% |
| Assay 1 used | 101,918 | 102,033 | 102,447 | 101,959 | 102,036 | 102,419 |
| Assay 2 used | 13,583 | 13,663 | 11,896 | 8,772 | 8,811 | 6,948 |
| Assay 3 used | 365 | 10,022 | 10,033 | 375 | 5,035 | 5,037 |
| Cost (US\$) | \$385,143 | \$409,869 | \$407,282 | \$374,144 | \$386,155 | \$383,216 |

|  | 1% positivity |  |  | 0.5% positivity |  |  |
| --- | --- | --- | --- | --- | --- | --- |
|  | 2-test | 3-test | WHO 2019 | 2-test | 3-test | WHO 2019 |
| HIV-negative classifications | 98,924 | 98,934 | 98,582 | 99,419 | 99,427 | 99,075 |
| HIV-positive classifications | 1035 | 979 | 971 | 542 | 490 | 486 |
| HIV-inconclusive | 41 | 87 | 446 | 40 | 83 | 439 |
| Observed positivity | 1.04% | 0.98% | 0.98% | 0.54% | 0.49% | 0.49% |
| False HIV-positive | 47.4 | 0.95 | 0.79 | 47.60 | 0.95 | 0.80 |
| False HIV-negative | 10.0 | 12.0 | 10.1 | 5.0 | 6.0 | 5.0 |
| PPV of entire testing strategy | 95.4% | 99.9% | 99.9% | 91.2% | 99.8% | 99.8% |
| NPV of entire testing strategy | >99.9% | >99.9% | >99.9% | >99.9% | >99.9% | >99.9% |
| Assay 1 used | 101,991 | 102,038 | 102,397 | 101,995 | 102,038 | 102,394 |
| Assay 2 used | 4,922 | 4,930 | 2,990 | 4,441 | 4,445 | 2,495 |
| Assay 3 used | 382 | 1,045 | 1,039 | 383 | 547 | 540 |
| Cost (US\$) | \$365,345 | \$367,184 | \$363,963 | \$364,245 | \$364,813 | \$361,556 |

**NPV:** negative predictive value; **PPV:** positive predictive value. **Note:** 2-test and 3-test denotes the number of consecutive HIV reactive tests to provide an HIV-positive diagnosis under the WHO 2015 guidelines 'high prevalence' and 'low prevalence' strategies, respectively, not number of tests required by the strategy. Specimens with repeated discrepant test results under the 2-test strategy proceed to a third assay. Specimens with repeat discrepant test results on the first two tests are ruled negative. In the 'WHO 2019' testing strategy, only A1 is repeated.

**Figure 2.**
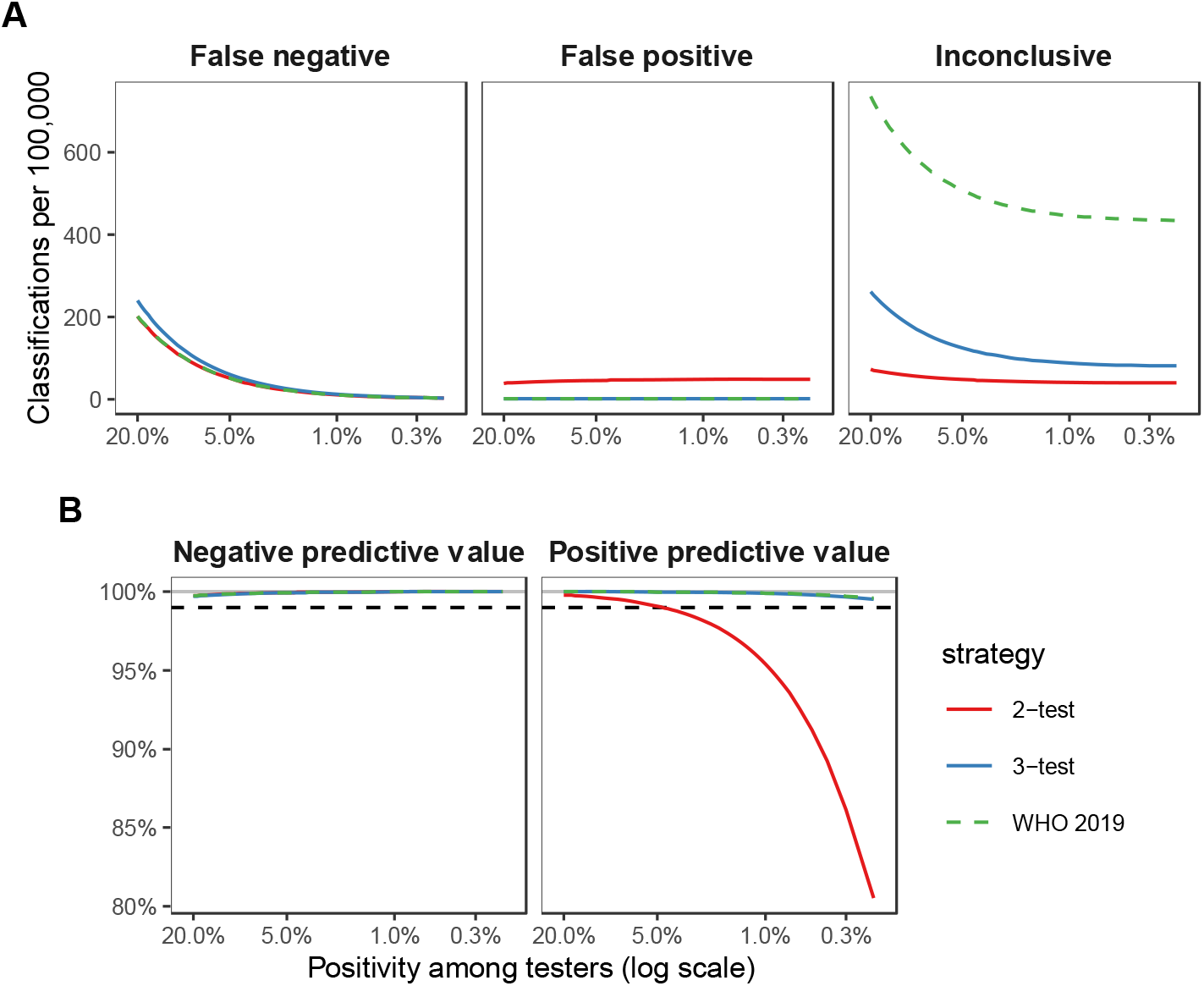
(A) Expected number of false negative, false positive, and inconclusive classifications per 100,000 clients tested under alternative testing strategies for true prevalence among HIV testing clients ranging from 20% to 0.2%. (B) Negative predictive value (NPV) and positive predictive value (PPV) for alternative testing strategies for prevalence ranging from 20% to 0.2%. Black dashed line indicates WHO target of 99% PPV and 99% NPV for HIV testing strategies.

The negative predictive value (NPV) was 99.9% or greater for all testing strategies at all positivity levels, well above the 99% threshold (Table 1, Figure 2B). Under the two-test strategy, the PPV was above the 99% threshold when positivity amongst individuals being tested was above 5%, but declined rapidly as positivity decreased (Table 1, Figure 2B). At 1% positivity, the PPV was 95% under the two-test strategy and this decreased to 91% at 0.5% positivity and 81% at 0.2% positivity. With the three-test or WHO 2019 testing strategy, the PPV increased to 99.9% at 1% positivity and was well above the 99% threshold for all simulated levels of positivity.

The number of results reported as ‘HIV-inconclusive’, in which the client would be asked to return for retesting after 14 days, was substantially higher under the WHO 2019 testing strategy than either the three-test or two-test strategy. For example, using the WHO 2019 testing strategy at 5% positivity, there were 508 (0.51%) HIV-inconclusive results per 100,000 individuals tested, compared to 124 per 100,000 (0.12%) for the three-test strategy and 47.2 (0.05%) with the two-test strategy. The increased number reported as HIV-inconclusive through the WHO 2019 testing strategy arose from two sources (Table 2). The majority arose when A1+ and A2– were discrepant, and the repeat A1 was HIV-reactive, which was reported as HIV-inconclusive under the WHO 2019 testing strategy. Under the WHO 2015 testing strategies, A2 was repeated and would result in either proceeding to A3 or reporting HIV-negative. Secondly, under the WHO 2019 testing strategy, cases that were reactive on each of the first two assays (A1+/A2+), then non-reactive on the third (A3–) were reported HIV-inconclusive. Under the two-test strategy, such individuals were reported HIV positive following A1+/A2+, and when positivity is below 5% the majority are false-positive misclassifications and are actually true negatives (Table 2).

**Table 2.** True HIV status for persons classified as ‘HIV-inconclusive’ under the WHO 2019 HIV testing strategy. All results per 100,000 persons tested.

|  | Testing Strategy Result Given |  |  | Positivity |  |  |  |
| --- | --- | --- | --- | --- | --- | --- | --- |
|  | 2-test | 3-test | WHO 2019 | 10% | 5% | 1% | 0.5% |
| Total inconclusive |  |  |  | 584 | 508 | 446 | 439 |
| True HIV-negative |  |  |  | 388 (66%) | 410 (81%) | 427 (96%) | 429 (98%) |
| True HIV-positive |  |  |  | 196 (34%) | 98 (19%) | 20 (4%) | 10 (2%) |
| Total A1+ / A2– / A1+ |  |  |  | 451 | 421 | 398 | 395 |
| True HIV-negative | Rep. A2 | Rep. A2 | INC | 353 (78%) | 372 (88%) | 388 (98%) | 390 (99%) |
| True HIV-positive | Rep. A2 | Rep. A2 | INC | 98 (22%) | 49 (12%) | 10 (2%) | 5 (1%) |
| Total A1+ / A2+ / A3– |  |  |  | 133 | 86 | 49 | 44 |
| True HIV-negative | False pos. | INC | INC | 35 (26%) | 37 (43%) | 39 (80%) | 39 (89%) |
| True HIV-positive | Corr. pos. | INC | INC | 98 (74%) | 49 (57%) | 10 (20%) | 5 (11%) |

**Rep. A2** = Under the WHO 2015 HIV testing strategies (2-test and 3-test), the A2 test is repeated.; **INC** = Inconclusive HIV test result returned with instruction to return after 14 days for retesting.; **False pos.** = A false positive HIV diagnosis is returned under the WHO 2015 2-test strategy; **Corr. pos.** = A correct HIV positive diagnosis is returned under the WHO 2015 2-test strategy.

### HIV testing commodities and programme costs

The number of A1 used per 100,000 tested was relatively similar and accounted for the majority of tests used for all positivity levels because most clients are classified HIV-negative following A1 and do not undergo further testing (Table 1, Figure 3A). The number of A2 required was the same under the three-test and two-test strategies and increased with positivity. The number of A2 was lower under the WHO 2019 testing strategy because A2 is not repeated following discrepant A1 and A2 results, which accounts for a substantial fraction of A2 usage under the WHO 2015 testing strategies when positivity is low. The number of A3 required was much greater for the three-test and WHO 2019 testing strategies, but even under these two testing strategy the number of A2 required were much larger than A3. This was because a substantial share of A2 are utilized on persons with discrepant A1+/A2– results, which are ultimately classified HIV-negative and do not progress on to A3. The difference in consumption between A2 and A3 increases as positivity declines.

**Figure 3.**
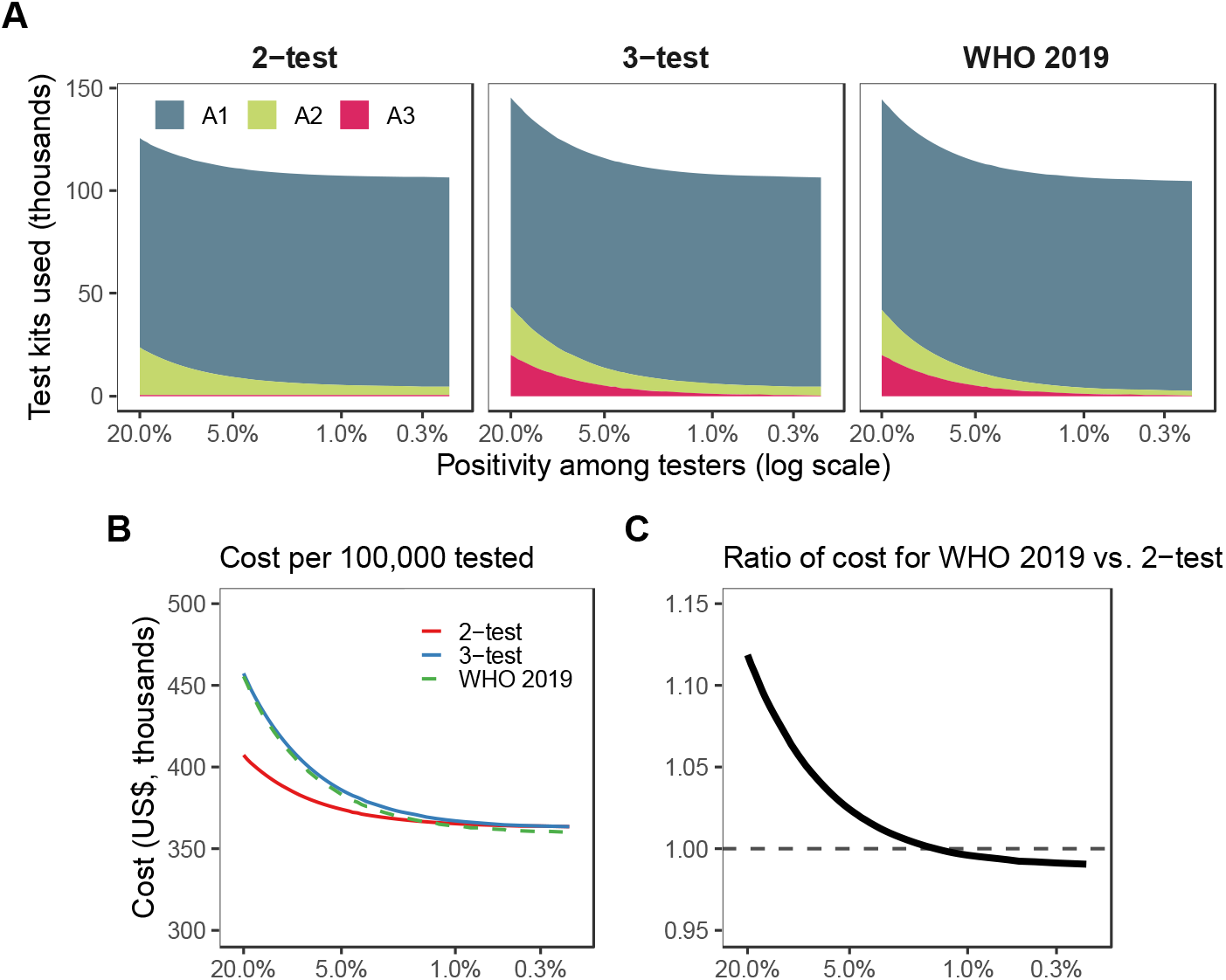
(A) Expected number of tests used by assay per 100,000 tested under the 2-test, 3-test, and WHO 2019 testing strategies for positivity ranging from 20% to 0.2%. (B) Estimated cost per 100,000 tested as a function of HIV positivity for each strategy. (C) Ratio of total cost under the WHO 2019 testing strategy versus the WHO 2015 2-test (high prevalence) strategy.

At 10% positivity, the total cost for the WHO 2019 testing strategy was 6% higher than the two-test strategy (Table 1; Figure 2C). The cost difference reduced considerably with declining positivity due to the lower consumption of A2 and A3. For testing positivity below 1.5%, the cost for the WHO 2019 testing strategy was slightly lower than the cost for two-test strategy because the reduced need for A2 outweighed the slight increase in A3 consumption.

### Estimated and projected changes in HTS outcomes: Malawi example

Figure 4 summarizes estimates and projections for the number of adult PLHIV over the period 2000 through 2025 in Malawi. The total adult PLHIV was projected to increase slightly through 2025 as PLHIV survive longer on ART. However, the proportion of undiagnosed PLHIV declined rapidly from an estimated 79% in 2005 to 33%, 10% and 5% in 2010, 2019 and 2025, respectively (Figure 4A). Consequently, despite large increases in the number tested for HIV each year, the number of HIV diagnoses peaked in 2010 and is projected to continue to decline (Figure 4B). This was reflected in rapidly declining positivity amongst clients tested. Before 2010, positivity in the testing program exceeded the population prevalence as symptomatic PLHIV were more likely to present for testing (Figure 4C). But in 2019, the positivity of individuals being tested for HIV declined to around 3.1% compared to adult HIV prevalence of 9.5% and the positivity was projected to further decline in 2025 to 1.2% compared to 8.3% prevalence. The proportion of individuals diagnosed for the first time among all tested for HIV was 1.2% in 2019, declining to 0.5% in 2025.

**Figure 4.**
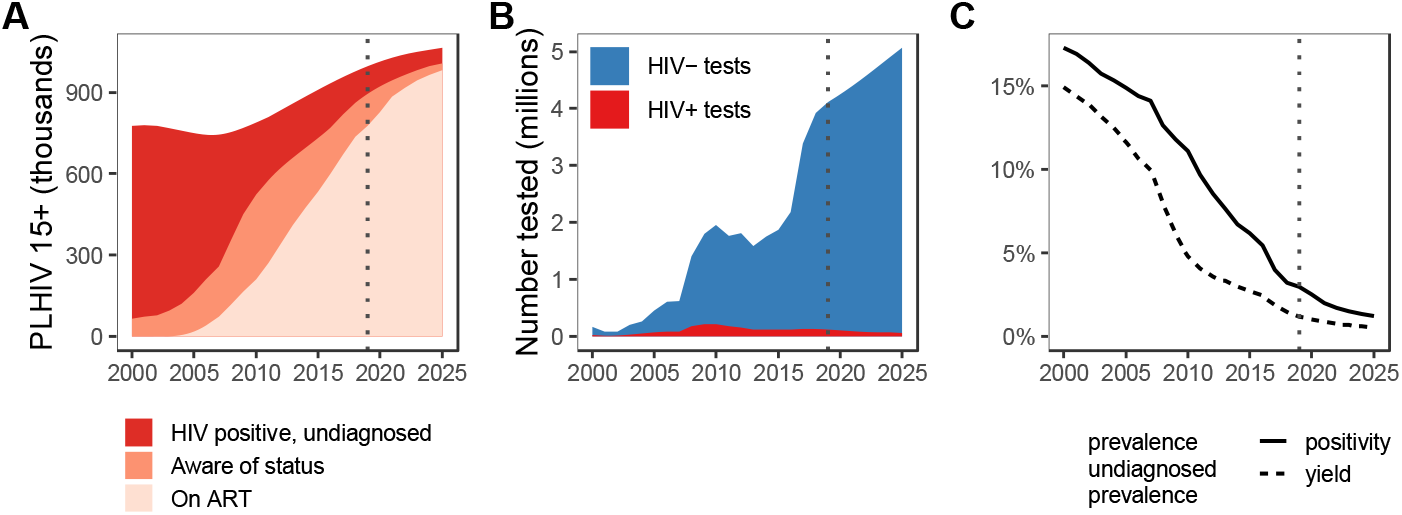
Estimates and projections for the HIV population and HIV testing programme in Malawi over period 2000 to 2025. (A) Total number of adult (15+ years) PLHIV stratified by undiagnosed, aware of HIV status but untreated, and on ART. (B) Total number of HIV tests per year amongst adults age 15+ and number of HIV-positive diagnoses. (C) HIV prevalence (light red shaded area) and prevalence of undiagnosed HIV (dark red) in the adult population (15+ years), and HIV positivity amongst those tested for HIV (solid line) and the ‘yield’ of new diagnoses (dotted line). ‘Positivity’ reflects the proportion of HTS clients who are positive including both new diagnoses and those re-testing who are already aware or on ART. ‘Yield’ of new diagnoses reflects the estimated proportion of HTS clients who being diagnosed for the first time. *(Source: Malawi 2020 UNAIDS Spectrum Estimates*.)

The consequences of declining positivity for expected HIV testing programme quality and costs are consistent with those reported in Table 1 and Figure 2. The expected NPV is substantially above the 99% threshold at all times (Figure 5A). However, the PPV dropped below the 99% threshold in 2018 with the WHO 2015 two-test strategy and will decline to 97% in 2025 if each assay performs at the minimum threshold required for WHO prequalification (Figure 5A). This equates to expecting an average of around 1800 false positive diagnoses per year. In contrast, the PPV remained above 99.9% with the WHO 2019 testing strategy—fewer than 40 false positive diagnoses per year. If the rates of HIV testing are maintained at current levels, an estimated 89,000 tests of A3 would be required to implement the WHO 2019 testing strategy in 2021, declining to 64,000 in 2025, compared to around 175,000 tests of A2 and over 4.5 million tests of A1 in 2021 (Figure 5B). The incremental cost of the WHO 2019 testing strategy was less than 2% in 2021, and declined to around 0.9% in 2025 (Figure 5C).

**Figure 5.**
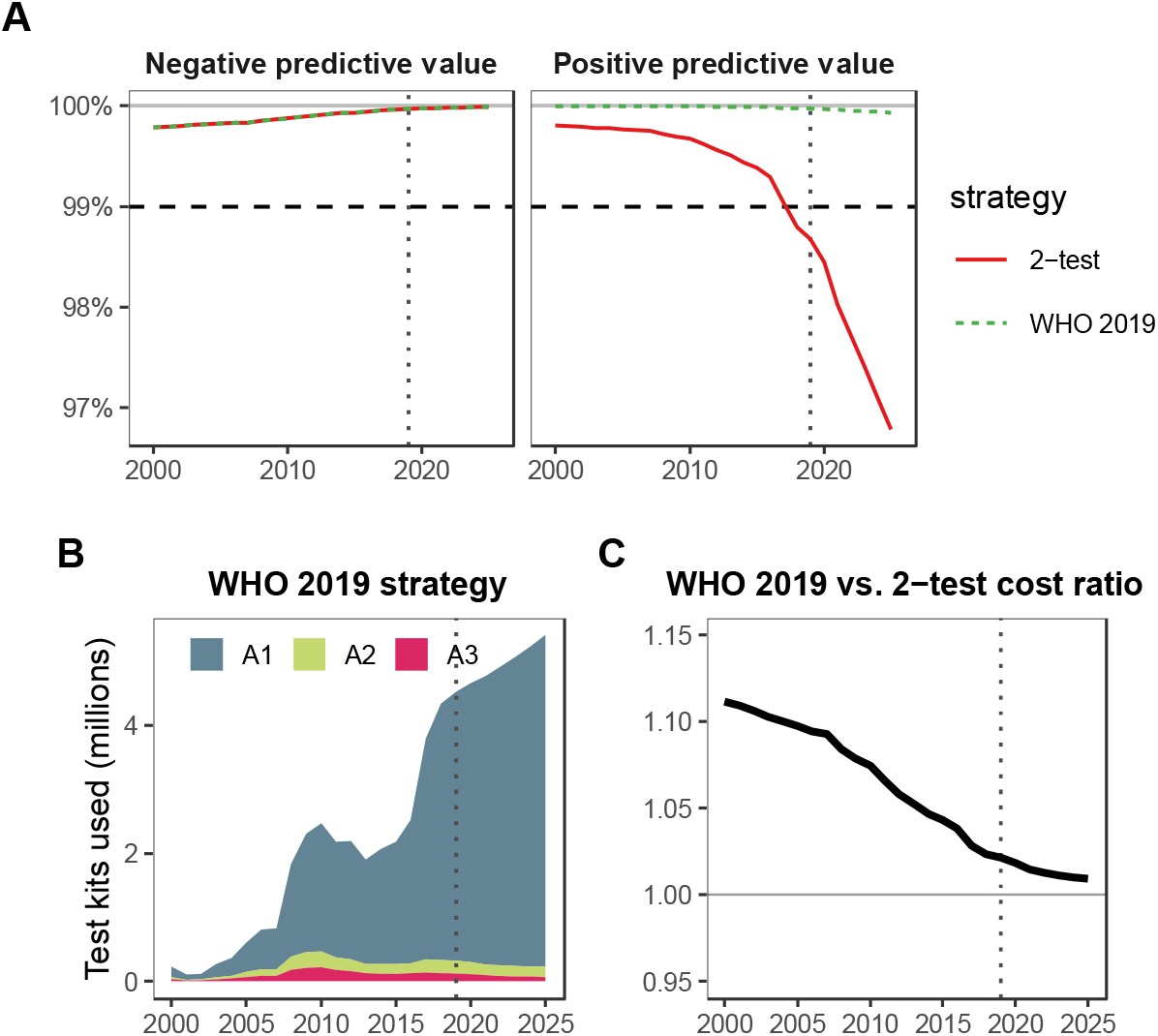
(A) Predicted negative predictive value (NPV) and positive predictive value (PPV) for adults presenting to HIV testing in Malawi over period 2000 to 2025 using the WHO 2015 ‘2-test’ (high prevalence) strategy compared to the WHO 2019 recommended strategy and assuming each assay performs at the minimum level required for WHO prequalification (99% sensitivity, 98% specificity). (B) Expected number of tests used by the WHO 2019 strategy. (C) Ratio of total cost per year for implementing the WHO 2019 strategy compared to the 2-test strategy.Dotted vertical line denotes year 2019 reflecting the final year of HIV testing programme data incorporated in estimates. Results for 2020 through 2025 reflect projections assuming HIV testing rates for 2019 continue through 2025.

## Discussion

Positivity in testing programs has declined below 5% even in countries with high HIV population prevalence [5]. The 2019 WHO HIV testing strategy requires reactive results on three serially conducted serology assays to diagnose HIV infection. This algorithm substantially reduced the number of false-positive misdiagnoses at all levels of positivity in HIV testing programs. When HIV assays perform at the minimum WHO pre-qualification threshold of 98% specificity, the 2019 WHO HIV testing strategy ensured that fewer than 1 in 100 persons diagnosed with HIV were false-positive, which was not the case for the two-test strategy used by many high prevalence countries.

The number of false-negative misdiagnoses reduced proportionally as positivity declined for HIV under all three testing strategies. The WHO 2019 testing strategy also increased the expected number of HIV-inconclusive results returned, for example from 0.04% under the two-test strategy to 0.45% with the WHO 2019 testing strategy at 1% positivity. However, at low positivity levels, increased HIV-inconclusive results arose primarily due to simplification of testing strategy for persons who are initially discrepant, reducing opportunity for errors, and among individuals who are truly HIV negative but would have been misclassified as HIV-positive under the two-test strategy. Had those HIV-inconclusive individuals received a false-positive misdiagnosis under the two-test testing strategy, the cost of misdiagnosis and life-long treatment would be significant [14,15].

The two-test and three-test testing strategies both require an A3 to be available, but the number of A3 consumed is expectedly much greater with the 3-test strategy since it is deployed for all individuals with reactive results for both A1 and A2 rather than only for those with discrepant test results (i.e. A1 reactive but A2 non-reactive). However, the number A3 required remains lower than A2 and the difference increases as positivity declines. This is because at low positivity, a large number of A2 are utilized for discrepant A1+/A2– results, which are classified as HIV negative by repeating A1 and do not proceed to A3.

Declining positivity amongst individuals who undergo HIV testing is a reflection of successful scale-up and implementation of HIV programmes that have reduced the undiagnosed HIV population [5]. Our case study of Malawi exemplifies the consequences for the HIV testing program. While HIV population prevalence has declined slowly from 11% in 2005 to 9.5% in 2019, the positivity amongst individuals undergoing HIV testing fell steeply from 15% to 3% over the same period. Assuming 98% specificity for each assay used, the 2015 WHO two-test testing strategy (designed for ‘high prevalence’ settings) was expected to perform below 99% PPV since 2018 in Malawi and a further decline in PPV was projected. To mitigate this, Malawi has pioneered quality testing initiatives, including verification testing (using the same two-test testing strategy in parallel) before ART initiation. Malawi service delivery data suggests steady improvements in testing accuracy which are reflected in a declining proportion of inconclusive results and discrepant verification testing results. While the true number of false-positive and false-negative misdiagnosis is unknown in Malawi, the simulated results illustrate the importance to consider the 2019 WHO testing strategy.

The incremental cost of switching to the WHO 2019 testing strategy was less than 2% of the total HIV testing programme cost and will decline further in the future. The low and decreasing incremental cost is because the large majority of HIV testing clients are HIV negative and do not proceed past A1, which thus accounts for the largest share of HTS programme cost despite a lower unit cost. Our analysis assumed that the incremental cost of switching to the WHO 2019 testing strategy was captured by the additional cost of delivering A3. However, in many settings A3 is not routinely stocked at all testing locations and specimens with discrepant test results are instead referred to laboratories for additional testing to confirm their HIV status. Additional training and supply chain costs of switching to a three-test strategy in such settings are uncertain. Incorporating a new A3 into the testing strategy would require training of all HTS providers, but such training activities could be conducted routinely as part of programme management, quality assurance, and implementation of other guideline changes. Resolving more cases at the facility instead of requiring discrepant specimens to be sent to a laboratory should also offset costs of transport, information management, and tracing clients to provide results. We were not able to estimate the cost for this additional lab testing using the current policy, but considerable delays in returning such results from the reference lab have highlighted the challenges with this approach. Timing changes and shifts in the testing strategy alongside other changes in updates to guidelines, log-books and integrated trainings can also increase cost efficiencies.

Consistent with WHO guidelines, we assumed that all tests in the algorithm were conducted serially, except for the parallel replication of A1 and A2 following initial discrepant A1+/A2–. An HIV diagnosis with the WHO 2019 testing strategy would require three finger pricks for clients with a reactive A1. There has been some consideration to conducting A2 and A3 in parallel could reduce the additional finger prick. However, this could also lead to additional challenges with the interpretation of discrepant test results and add substantial wastage of A3 since, as positivity becomes lower, an increasing share of those progressing to A2 were false reactive to A1 and classified as HIV negative on A1 repetition, and not progress to A3. For example, in Malawi, parallel application of A2 and A3 would roughly double the number of A3 required.

There were two assumptions with limited empirical data to which our findings are potentially sensitive: (1) that 80% of discrepant A1+/A2– would be correctly resolved upon replication of the A1 and A2, while 20% would remain discrepant, and (2) that each assay in the testing strategy is independent and performs at the minimum threshold for WHO prequalification, namely 98% specificity. Our conclusions should be interpreted considering these limitations and should be reevaluated as more evidence on these points becomes available.

Our modelling suggests that transitioning to the WHO 2019 HIV testing strategy, requiring three reactive tests to diagnose HIV, ensures accuracy and quality of HIV diagnosis into the future with minimal consequences for overall HIV testing programme costs. The alternative, without adopting the strategy requiring three reactive tests to diagnose HIV, is increasing false positive misdiagnoses as a proportion of all those newly diagnosed with HIV. This incurs unnecessary lifelong ART costs for HIV negative persons [14,15], unquantified individual physical, psychosocial and emotional harms of receiving a false positive HIV misdiagnosis [16,17], and undermining confidence in the health system. While the WHO 2019 HIV testing strategy ensures the fidelity of HIV diagnosis and subsequent care and treatment, the main determinant of HIV testing programme cost is the overall scale if HIV testing and the number A1 conducted.

## Data Availability

The national Spectrum file for Malawi are available upon request from UNAIDS: https://www.unaids.org/en/dataanalysis/datatools/spectrum-epp

## References

1. Global programme on AIDS. Recommendations for the selection and use of HIV antibody tests. Wkly Epidemiol Rec Relev Epidemiol Hebd World Heal Organ. 1992;67:145–149.

2. Joint United Nations Programme on HIV/AIDS (UNAIDS)-WHO. Revised recommendations for the selection and use of HIV antibody tests. Wkly Epidemiol Rec. 1997;72:81–7. Available: http://www.ncbi.nlm.nih.gov/pubmed/9238418

3. World Health Organization. Selecting and purchasing HIV, HBsAg and HCV in vitro diagnostics. 2021 [cited 23 Feb 2021]. Available: https://www.who.int/diagnostics_laboratory/procurement/purchase/en/

4. World Health Organization. Consolidated guidelines on HIV testing services. 2015 p. 193. Available: http://apps.who.int/iris/bitstream/10665/179870/1/9789241508926_eng.pdf

5. Giguère K, Eaton JW, Marsh K, Johnson LF, Johnson CC, Ehui E, et al. Trends in knowledge of HIV status and efficiency of HIV testing services in sub-Saharan Africa, 2000–20:a modelling study using survey and HIV testing programme data. Lancet HIV.2021 [cited 8 Mar 2021]. doi:10.1016/S2352-3018(20)30315-5

6. Fonner VA, Sands A, Figueroa C, Baggaley R, Quinn C, Jamil MS, et al. Country adherence to WHO recommendations to improve the quality of HIV diagnosis: a global policy review. BMJ Glob Heal. 2020;5:e001939. doi:10.1136/bmjgh-2019-001939

7. World Health Organization. Consolidated guidelines on HIV testing services 2019. 2019 [cited 23 Feb 2021] p. 261. Available: https://apps.who.int/iris/rest/bitstreams/1313903/retrieve

8. WHO Prequalifications Team: Diagnostics. Technical specification series for submission to WHO prequalification – Diagnostic Assessment: TSS-1: Human Immunodeficiency Virus (HIV) rapid diagnostic tests for professional and/or self-testing. 2016 [cited 15 Nov 2018]. Available: http://apps.who.int/iris/bitstream/handle/10665/251857/9789241511742-eng.pdf

9. World Health Organization. Global Price Reporting Mechnism. 2017 [cited 10 Dec 2017]. Available: https://apps.who.int/hiv/amds/price/hdd

10. Government of Malawi Ministry of Health. Integrated HIV Program Report October-December 2017. Lilongwe; 2018.

11. Stover J, Glaubius R, Mofenson L, Dugdale CM, Davies MA, Patten G, et al. Updates to the Spectrum/AIM model for estimating key HIV indicators at national and subnational levels. AIDS. 2019;33:S227–S234. doi:10.1097/QAD.0000000000002357

12. Eaton JW, Brown T, Puckett R, Glaubius R, Mutai K, Bao L, et al. The Estimation and Projection Package Age-Sex Model and the r-hybrid model: new tools for estimating HIV incidence trends in sub-Saharan Africa. AIDS. 2019;33 Suppl 3: S235–S244. doi:10.1097/QAD.0000000000002437

13. Maheu-Giroux M, Marsh K, Doyle CM, Godin A, Lanièce Delaunay C, Johnson LF, et al. National HIV testing and diagnosis coverage in sub-Saharan Africa. AIDS. 2019;33:S255–S269. doi:10.1097/QAD.0000000000002386

14. Eaton JW, Johnson CC, Gregson S. The Cost of Not Retesting: Human Immunodeficiency Virus Misdiagnosis in the Antiretroviral Therapy “Test-and-Offer” Era. Clin Infect Dis. 2017;65. doi:10.1093/cid/cix341

15. Lasry A, Kalou MB, Young PR, Rurangirwa J, Parekh B, Behel S. Cost implications of HIV retesting for verification in Africa. PLoS One. 2019;14. doi:10.1371/journal.pone.0218936

16. Bhattacharya R, Barton S, Catalan J. When good news is bad news: Psychological impact of false positive diagnosis of HIV. AIDS Care -Psychol Socio-Medical Asp AIDS/HIV. 2008;20:560–564. doi:10.1080/09540120701867206

17. Johnson CC, Dalal S, Baggaley R, Taegtmeyer M. A public health approach to addressing and preventing misdiagnosis in the scale-up of HIV rapid testing programmes. Journal of the International AIDS Society. International AIDS Society; 2017. doi:10.7448/IAS.20.7.22190

